# Rotational worker vaccination provides indirect protection to vulnerable groups in regions with low COVID-19 prevalence

**DOI:** 10.1101/2021.10.05.21264557

**Authors:** Maria M. Martignoni, Proton Rahman, Amy Hurford

## Abstract

As COVID-19 vaccines become available, different model-based approach have been developed to evaluate strategic priorities for vaccine allocation to reduce severe illness. One strategy is to directly prioritize groups that are likely to experience medical complications due to COVID-19, such as older adults. A second strategy is to limit community spread by reducing importations, for example by vaccinating members of the mobile labour force, such as rotational workers. This second strategy may be appropriate for regions with low disease prevalence, where importations are a substantial fraction of all cases and reducing the importation rate reduces the risk of community outbreaks, which can provide significant indirect protection for vulnerable individuals. Current studies have focused on comparing vaccination strategies in the absence of importations, and have not considered allocating vaccines to reduce the importation rate. Here, we provide an analytical criteria to compare the reduction in the risk of hospitalization and intensive care unit (ICU) admission over four months when either older adults or rotational workers are prioritized for vaccination. Vaccinating rotational workers (assumed to be 6,000 individuals and about 1% of the Newfoundland and Labrador (NL) population) could reduce the average risk of hospitalization and ICU admission by 42%, if no community spread is observed at the time of vaccination, because epidemic spread is reduced and vulnerable individuals are indirectly protected. In contrast, vaccinating all individuals aged 75 and older (about 43,300 individuals, or 8% of the NL population) would lead to a 24% reduction in the average risk of hospitalization, and to a 45% reduction in the average risk of ICU admission, because a large number of individuals at high risk from COVID-19 are now vaccinated. Therefore, reducing the risk of hospitalization and ICU admission of the susceptible population by reducing case importations would require a significantly lower number of vaccines. Benefits of vaccinating rotational workers decrease with increasing infection prevalence in the community. Prioritizing members of the mobile labour force should be considered as an efficient strategy to indirectly protect vulnerable groups from COVID-19 exposure in regions with low disease prevalence.

## Introduction

Vaccines for COVID-19 have become available (Corum et al., 2020), and authorities are facing decisions as to which groups to prioritize to mitigate severe illness due to COVID-19. Model-based approaches have been developed to compare the efficiency of different vaccination strategies in reducing hospitalization, intense care unit (ICU) admission, death, or long COVID (Bubar et al., 2021; Mulberry et al., 2021; Chen et al., 2021). A common strategy has been to prioritize vulnerable individuals, defined as people at high risk from COVID-19 (NHS), such as older adults (Bubar et al., 2021), or people with health conditions (e.g., heart or lung conditions, weakened immune systems, obesity, or diabetes) (Persad et al., 2021). Other effective vaccination strategies have included prioritizing essential workers (Mulberry et al., 2021), or individuals with a large number of social contacts (Chen et al., 2021).

An alternative approach is to vaccinate groups that are likely to introduce the disease into the community, such as rotational workers or other members of the mobile labour force (Lemke, 2021). These are workers that, to perform essential functions, are required to cross provincial or international borders on a regular basis (Neis et al., 2020). Examples include fisheries workers, truck drivers, flight crews, or other individuals that alternate times away at work with time at home. These workers may become infected with COVID-19 while working in another province, and when returning home may initiate a community outbreak. No current studies have considered how reducing the importation rate by vaccinating rotational workers could lower the risk of community infections and severe illness due to COVID-19 by preventing spread to all individuals, including those in vulnerable groups.

The population of Newfoundland and Labrador (NL) is ∼522,000 and more than 90% of the population lives on the island of Newfoundland (Statistica, 2021). The island has very few ports of entries and during the pandemic has been subject to stringent border control (Hurford et al., 2021). In part, due to these strict measures, NL has experienced very minimal community spread for extended periods of time (Berry et al., 2020). In NL rotational workers are a significant part of the work force (Hewitt et al., 2018), where a rotational worker is defined as ‘a resident of Newfoundland and Labrador who travels to another province or territory of Canada to work, on a set schedule of time away at work alternating with time at home in Newfoundland and Labrador’ (GovNL, 2021). The large majority of rotational workers from NL are intraprovincial workers employed in Ontario, Alberta, and Nova Scotia (Hewitt et al., 2018), and since October 2020 infection prevalence in Ontario and Alberta has been substantially higher than in NL (Berry et al., 2020). Vaccinating rotational workers could reduce the rate at which the virus is imported in a region, reducing the chance of a community outbreak and providing indirect protection for those individuals that are likely to experience medical complications due to COVID-19.

Here, we quantify the reduction in the risk of hospitalization and ICU admission if: (1) vulnerable groups in the community are vaccinated, thus reducing the average probabilities of hospitalization and ICU admission across the remaining susceptible population, where the average is calculated by considering the age structure and the probabilities of hospitalization and ICU admission for each age group in the susceptible population (see Appendix A); or (2) rotational workers are vaccinated, thus reducing the rate that the virus is imported in the community, and, in turn, reducing the risk of community outbreaks, but leaving probabilities of hospitalization and ICU admission of the susceptible population unchanged. We derive a concise analytical criteria to compare the reduction in risk occurring when strategies (1) and (2) are applied. The criteria is based on easily accessible quantities, such as infection prevalence in the community at the time of vaccination, the number of importations to the province due to non-self-isolating rotational workers, the percent reduction in viral transmission after vaccination, and the probabilities of hospitalization and ICU admission of the vulnerable group vaccinated (Fig. 1). We calculate the risk of hospitalization and ICU admission for NL when rotational workers or older adults are vaccinated. Our analysis provides additional support for public health decisions regarding vaccine prioritization strategies.

**Fig. 1:**
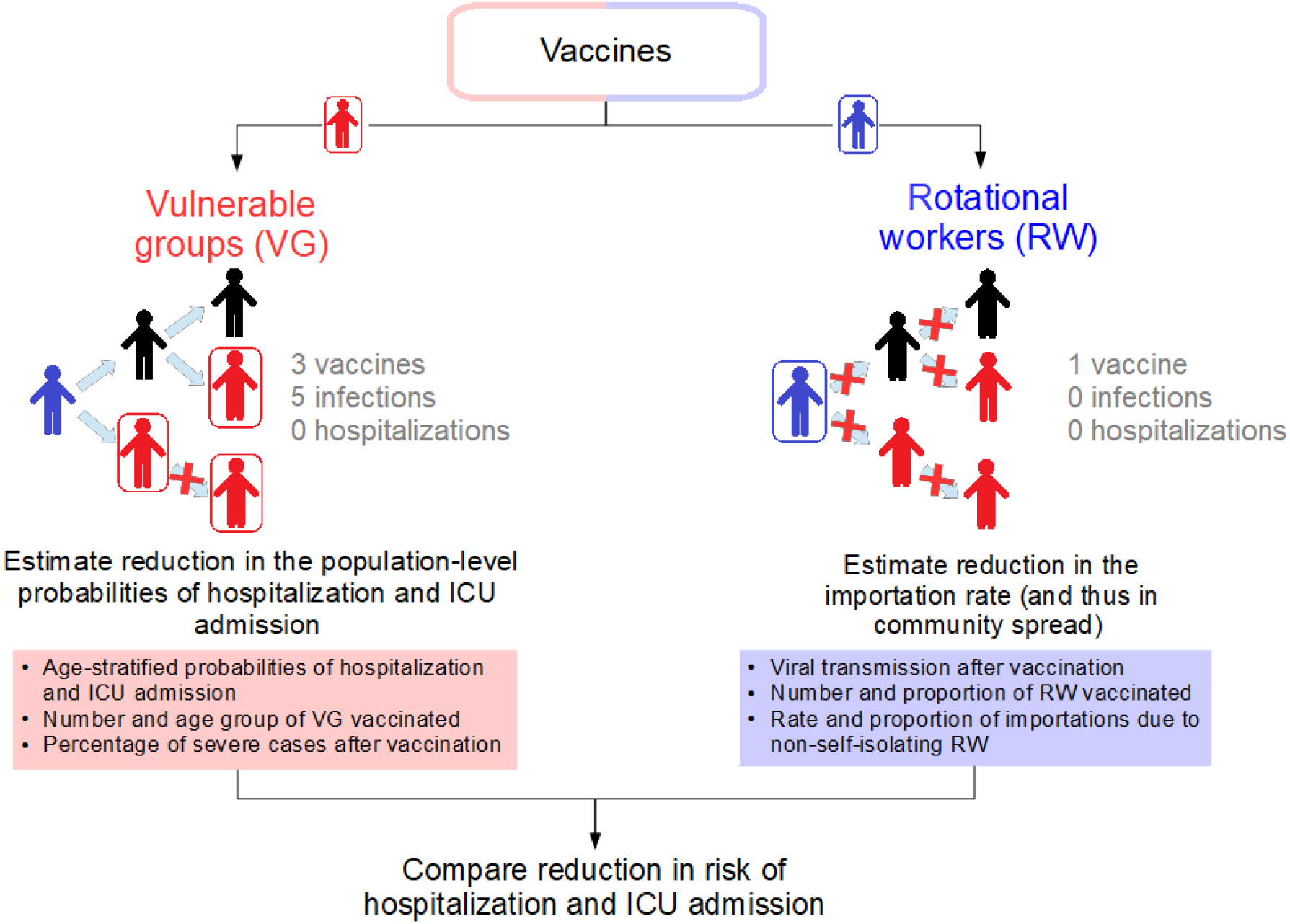
Schematic representation of our research question. Our objective is to quantify the reduction in the risk of hospitalization and ICU admission when vulnerable groups (red individuals in the figure) or rotational workers (blue individuals) are vaccinated. Black individuals represent other non-vulnerable community members. A box around an individual symbolizes vaccination. Vaccinating vulnerable groups reduces the average probabilities of hospitalization and ICU admission for the remaining susceptible population. Vaccinating rotational workers reduces the importation rate, and prevents community outbreaks that may otherwise have occurred.

## Model and Methods

### Modelling the impact of vaccination on the infection dynamics

We use a SIR model (Kermack and McKendrick, 1927) with importations to simulate changes in the infectious status of a population as a consequence of community and disease introduction from external sources. The disease can spread from infected individuals *I* to susceptible individuals *S*, with a transmission rate *β* per day. The rate that infected individuals enter the province and fail to self-isolate is *m* individuals per day, and all infected individuals are assumed to be immediately infectious. Infected individuals recover from infection (or die) at rate *γ* per day, and are subsequently removed from the description of the infection dynamics. We assume that the rate that individuals enter the province remains low over time, and the total population size can be approximated by the constant *N*. The system of differential equation representing the dynamics described above is:

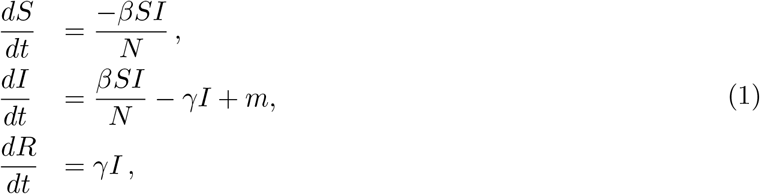

with initial conditions {*S*_0_, *I*_0_, *R*_0_}. We assume that at time zero a certain number of individuals in the community(*V*_0_ ≥ 0) are already vaccinated. Additionally, we assume that at this time a certain number of vaccines are available to vaccinate either vulnerable individuals (*V*_*g*_), or rotational workers *V*_*w*_. Therefore we will consider the situations where the same number *V* of either vulnerable individuals or rotational workers are vaccinated (i.e., either *V*_*g*_ = *V* and *V*_*w*_ = 0, or *V*_*w*_ = *V* and *V*_*g*_ = 0), as well as the situation where neither rotational workers nor vulnerable individuals are vaccinated (*V*_*g*_ = *V*_*w*_ = 0). We assume that vaccination reduces the probability that susceptible individuals become infected and transmit the disease by (1 − *Z*), where *Z* = 0 indicates that vaccination completely prevents viral transmission, while *Z* = 1 indicates that vaccination does not prevent viral transmission. We derive the following initial condition for the number of susceptible individuals in the community at time zero:

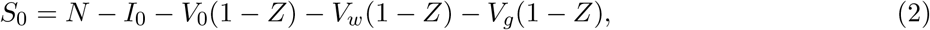

where *I*_0_ represents infection prevalence at time zero, and is understood as the number of cases that are not in self-isolation and are spreading infections undetected in the community at the time vaccination begins.

Unvaccinated rotational workers are part of the susceptible class, but can also introduce the disease into the community through importations. The importation rate (*m* infections per day) is determined by the total number of rotational workers *W*, by the number of rotational workers vaccinated (*V*_*w*_), by the proportion of importations due to non-self isolating rotational workers (*η*), by the importation rate due to other non-self-isolating travelers (*ω* individuals per day), and by the reduction in viral transmission after vaccination (*Z*). As such:

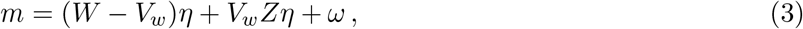

and vaccinating rotational workers reduces the importation rate *m*. We define *m*_0_ as the importation rate before rotational workers are vaccinated, and *m*_*r*_ as the reduced importation rate when rotational workers have been vaccinated. Vaccinating vulnerable groups does not affect the importation rate. Model parameters and default values used for the simulations are summarized in Table 1.

**Table 1:** Description of model variables and parameters used for the simulations. Default values are based on population structure and epidemiological features of NL.

| Symbol | Description | Default value |
| --- | --- | --- |
| $S$ | Susceptible individuals | — |
| $I$ | Infected individuals (not in self-isolation) | — |
| $R$ | Recovered individuals | — |
| $S_0$ | Susceptible individuals at time zero | see Eq. (2) |
| $I_0$ | Infected individuals (not in self-isolation) at time zero | 0 to 60 individuals |
| $N$ | Total population | 522,103 individuals (Statistica, 2021) |
| $T$ | Time frame considered for the simulations | 120 days |
| $C(T)$ | Number of cumulative cases over a time interval T | — |
| $\beta$ | Transmission rate | $1.1\gamma$ per day * |
| $\beta_0$ and $\beta_v$ | Transmission rates before and after vaccination | $\beta S_0/N$ |
| $\gamma$ | Recovery rate | 1/14 per day (Hurford and Watmough, 2021) |
| $m_0$ and $m_r$ | Importation rates before and after vaccinating RW | — |
| $Z$ | Probability of viral transmission after vaccination | 0.67 (Wise, 2021) |
| $W$ | Number of rotational workers | 6,000 individuals * |
| $\eta$ | Rate of imported cases per non-self-isolating RW | 0.4/W to 2/W per month * |
| $\omega$ | Rate of imported cases due to other non-self-isolating travelers | 0.4 to 2 per month * |
| $V_0$ | Number of individuals vaccinated before time zero | ~10,000 individuals ** |
| $V_g$ and $V_w$ | Number of VG or RW vaccinated at time zero | ~6'000 individuals ** |
| $P_h$ and $P_{h_r}$ | Average probabilities of hospitalization before and after vaccinating VG | 7.89% and 7.61% ** |
| $P_u$ and $P_{u_r}$ | Average probabilities of ICU admission before and after vaccinating VG | 2.35% and 2.14% ** |

\* Estimated parameters. See Appendix B for variations on the default values. \*\* See Appendix A for derivation.

### Quantification of risk reduction due to vaccination

To quantify the reduction in the risk of hospitalization and ICU admission of a community when rotational workers (RW) or vulnerable groups (VG) are vaccinated, we define:

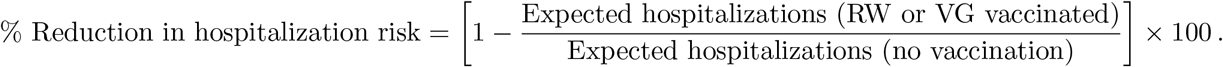

An equivalent equation can be written for the % reduction in the risk of ICU admission.

To calculate the expected number of hospitalizations and ICU admission we compute the number of cumulative infections *C*(*T*) over a certain period of time *T* by using Eq. (1). We multiply then *C*(*T*) by the average probabilities of hospitalization (*P*_*h*_) or ICU admission (*P*_*u*_) of the susceptible population and obtain:

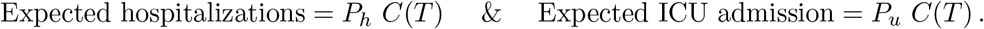

Vaccinating RW reduces the expected number of hospitalizations by reducing the importation rate (see Eq. (3)), and thus the cumulative number of infectious cases. Vaccinating vulnerable groups reduces the average probabilities of hospitalization (from *P*_*h*_ to 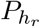) and ICU admission (from *P*_*u*_ to 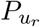) for the susceptible population because vaccinated vulnerable individuals are no longer included in the calculation of this average. (see Table 1 and Appendix A).

### Application to Newfoundland and Labrador

We compare the reduction in the risk of hospitalization and ICU admission in NL when vaccinating either rotational workers or vulnerable individuals. We estimate that NL has about 6,000 rotational workers. The census data (Population and Demographics) indicates that in NL about 12,000 individuals are between 80 and 84 years old. We consider half of the older adults in the 80-84 age group to be the vulnerable individuals of interest for our analysis, as this group offers a good size comparison with RW. Additionally, we consider that individuals aged 85 or older have already received the vaccine. The same analysis could be performed for a different vulnerable group, whose vaccination leads to a decrease in the average probabilities of hospitalization and ICU admission of the susceptible population.

Using the NL census data (Population and Demographics), the age-stratified probabilities of hospitalization and ICU admission (Ferguson et al., 2020; Kronbichler et al., 2020), and the distribution of asymptomatic cases by age groups (Kronbichler et al., 2020), we calculate the average probabilities of hospitalization and ICU admission in NL, where the average is taken across the age-structure in the susceptible population. The exact data used are given in Appendix A. Our estimates are *P*_*h*_ = 7.89% for the average probability of hospitalization in NL due to COVID-19 infection, and *P*_*u*_ = 2.35% for the average probability of ICU admission (if individuals aged 85 and above are vaccinated). Assuming that vaccination prevents severe illness (Voysey et al., 2021; Knoll and Wonodi, 2021), vaccinating 6,000 older adults aged 80-84 would lower the average probability of hospitalization of the remaining susceptible population to 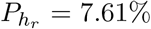, and the average probability of ICU admission to 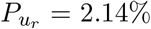. We estimate that NL may experience 0.4 to 2 importations of cases of non-self-isolating individuals every month, and we assume rotational workers are responsible for 20% to 80% of those importations.

## Results

### Quantification of risk reduction due to vaccination

The risk of hospitalization when vulnerable groups are vaccinated can be expressed as

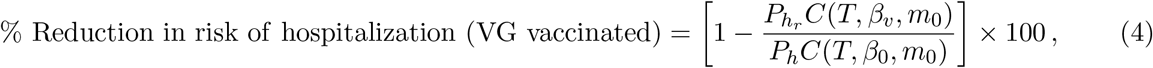

where *P*_*h*_ and 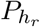 are the average probabilities of hospitalization before and after vaccination and *C*(*T, β, m*) is the number of cumulative cases, which depends on the time interval *T* considered, on the transmission rate before or after vaccination (*β*_*j*_ = *βS*_0_*/N*, for *j* = 0, *v*), and on the importation rate *m*_0_. Similarly, the reduction in risk of ICU admission when VG are vaccinated is given by

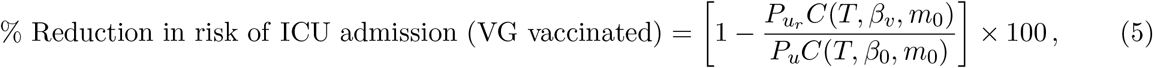

where *P*_*u*_ and 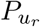 are the average probabilities of ICU admission before and after vaccination.

The reduction in the risk of hospitalization and ICU admission when RW are vaccinated is:

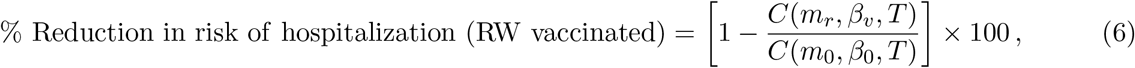

where *m*_0_ and *m*_*r*_ represent the importation rates before and after vaccination, and depend on the number of RW (*W*), on the number of RW vaccinated (*V*_*w*_), on viral transmission after vaccination (*Z*), on the proportion of successful importations due to non-self-isolating rotational workers (*η*), and on the importation rate due to other non-self-isolating travelers (*ω*) (see Eq. (3)). Note that the reduction in the risk of hospitalization when RW are vaccinated is equal to the reduction in the risk of ICU admission, as the probabilities *P*_*h*_ and *P*_*u*_ remain unchanged before and after vaccination of RW, and can be canceled from the numerator and the denominator of the fraction of Eq. (6).

We can determine when a reduction in the risk of hospitalization due to vaccinating VG equals a reduction in the risk of hospitalization obtained by vaccinating RW as follows:

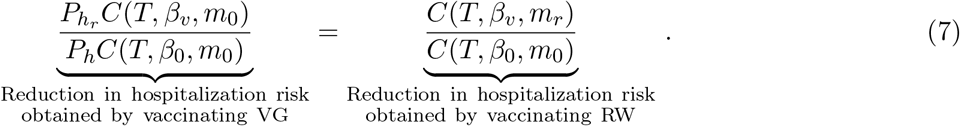

Eq. (7) can be simplified as:

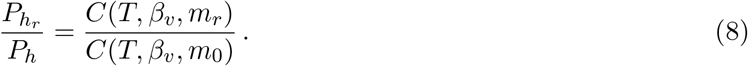

The risk of hospitalization can therefore be equally reduced by a reduction in the average probability of hospitalization, or by a reduction in the expected number of cumulative infections.

When infection prevalence remains low, and changes in the susceptible population over a short period of time remain small, the system of Eq. (1) can be approximated by a single linear differential equation in *I*, namely:

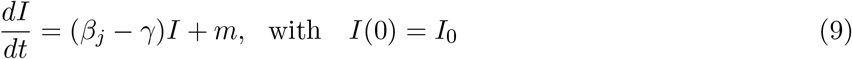

with solution

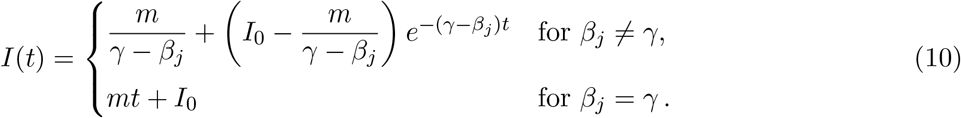

where *β*_*j*_ = *βS*_0_*/N*, for *j* = 0, *v*. The cumulative number of infections can be computed by solving the integral:

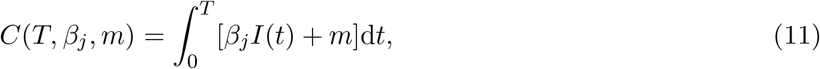

with analytical solution

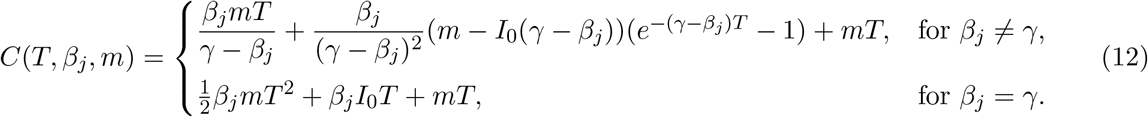

Note that if infection prevalence is initially zero (i.e., *I*_0_ = 0), the number of cumulative cases is linearly proportional to the importation rate *m* (cfr. Eq. (12); to see this note that *C*(*m, β*_*j*_, *T*) = *mC*(*β*_*j*_, *T*) for *I*_0_ = 0), and Eq. (8) can be rewritten as

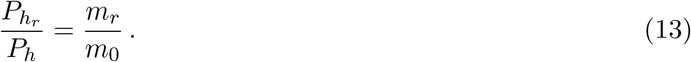

Thus, for *I*_0_ = 0 (i.e., in the absence of community spread at vaccination time), an *X*% reduction in the importation rate has the same impact on reducing the risk of hospitalization as an *X*% reduction in the average probability of hospitalization. Note that Eq. (13) does not depend on the transmission rate *β* in the community or on the time interval *T* considered.

In contrast, for *I*_0_ > 0, an *X*% reduction in the probability of hospitalization has a much larger impact than the same *X*% reduction in the importation rate. For example, if *I*_0_ = 20, a 70% reduction in the importation rate corresponds to a 10% reduction in the probability of hospitalization. A graphical representation of Eq. (8) as a function of infection prevalence at the initial time is given in Fig. 2. The importance of vaccinating rotational workers when *I*_0_ > 0 decreases more rapidly when the importation rate is low, when viral transmission is high and when shorter time intervals are considered (see Appendix B, Figs. B.1 and B.2). An analogous relationship to Eq. (8) can be written to compare a reduction in the probability of ICU admission and a reduction in the importation rate:

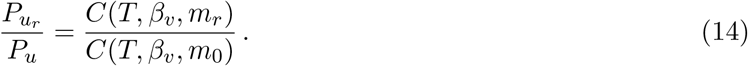

**Fig. 2:**
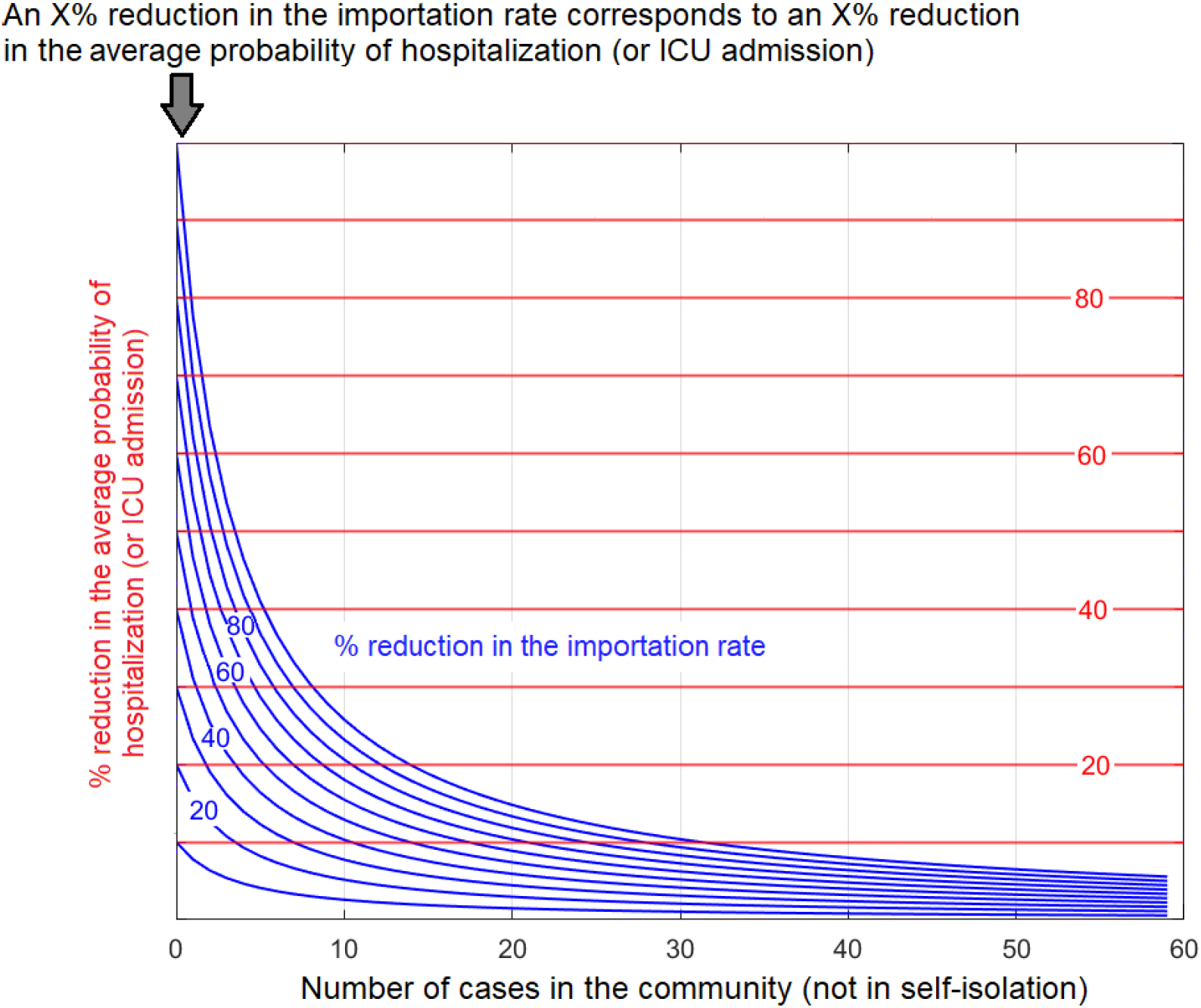
Equivalence between an X% reduction in the importation rate (blue lines) and an X% reduction in the average probability of hospitalization (red lines) in reducing the risk of hospitalization, as a function of the number of infectious cases in the community *I*_0_ (not in self-isolation) (cfr. Eq. (8)). A reduction in the importation rate can be achieved by vaccinating rotational workers. A reduction in the average probability of hospitalization can be achieved by vaccinating vulnerable groups in the community. Note that in the absence of community spread (i.e., *I*_0_ = 0) an X% reduction in the importation rate corresponds to an X% reduction in the average probability of hospitalization for the susceptible population (cfr. Eq. (13)). When infection prevalence is larger than zero, a reduction in the importation rate has a smaller impact than the same relative reduction in the average probability of hospitalization. For example, if *I*_0_ = 20, a 70% reduction in the importation rate corresponds to a 10% reduction in the average probability of hospitalization. Also shown is the equivalence between a reduction in the importation rate and the average probability of ICU admission (cfr. Eq. (14)).

### Application to Newfoundland and Labrador

Fig. 3 shows the reduction in the risk of hospitalization obtained when 6,000 rotational workers (corresponding to 1% of the total population) or 6,000 individuals between 80 and 84 years (i.e., half of the 80-84 age group in NL) are vaccinated. We can see that if infection prevalence is initially zero and if rotational workers are responsible for 60% of the importations, vaccinating rotational workers decreases the risk of hospitalization and ICU admission by 42%. Alternatively, vaccinating older adults would lead to a 7% reduction in the risk of hospitalization and a 12% reduction in the risk of ICU admission for the susceptible population. Even if non-self-isolating rotational workers are responsible for only 20% of importations, vaccination would lead to a 16% reduction in the risk of hospitalization and ICU admission, which is still higher than the reduction achieved by vaccinating half of the 80-84 age group.

**Fig. 3:**
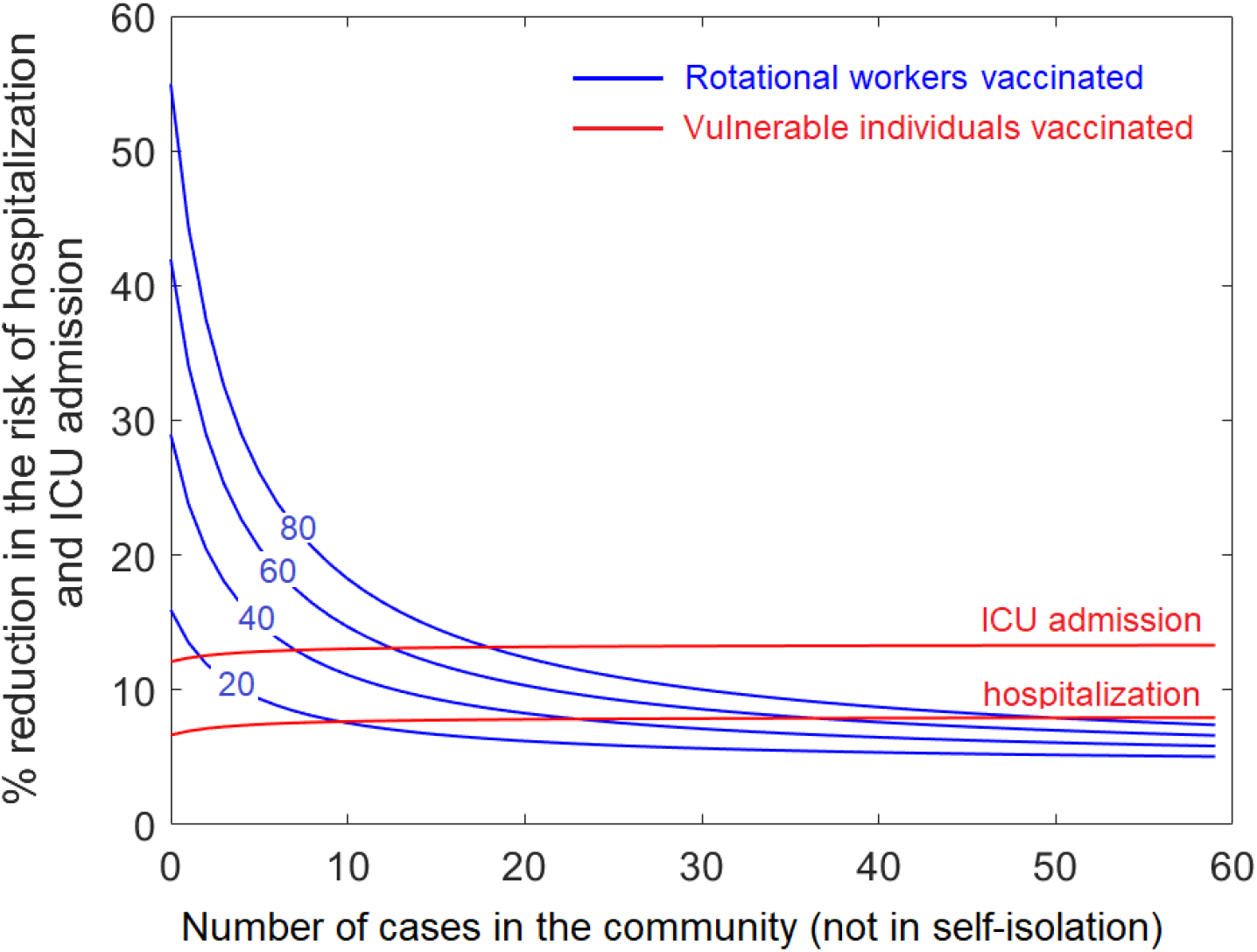
Percent reduction in the risk of hospitalization and ICU admission when rotational workers (blue curves) or half of adults in the 80-84 age group in NL are vaccinated (red curves). The numbers on the blue lines are the percentage of importations due to non-self-isolating rotational workers. Vaccinating rotational workers decreases the risk of hospitalizations and ICU admissions alike (see Eqs. (6)). Results are obtained using the model presented in the system of Eq. (1), with parameter values given in Table 1.

Using Eqs. (3) and (13) we can show that, if infection prevalence at the time of vaccination is zero, vaccinating rotational workers may ensure a large reduction in the risk of hospitalization and ICU admission with a lower number of vaccines. We estimate that if vaccination reduces viral transmission by 67% (Wise, 2021), and if rotational workers are responsible of 60% of the importations (with 1.2 importations per month due to non-self-isolating rotational workers and 0.8 non-self-isolated importations per month due to other sources, i.e., *Wη* = 1.2/30 and *ω* = 0.8/30), vaccinating 6,000 rotational workers would lead to a 42% reduction in the importation rate, and thus to an equivalent reduction in the risk of hospitalization and ICU admission (see Eq. (13) and Fig. 3). In contrast, vaccinating all individuals aged 75 and older in NL (about 43,300 individuals, or 8% of the total population) would lead to only a 24% reduction in the risk of hospitalization, and a 45% reduction in the risk of ICU admission.

The benefits of vaccinating rotational workers decrease rapidly with increasing number of infectious cases in the community. With 50 infectious cases in the community (not in self-isolation), vaccinating half of the 80-84 age group has a nearly equivalent impact on reducing the risk of hospitalization as vaccinating rotational workers, even if rotational workers are responsible for 80% of the imported cases. With only 20 infectious cases, the reduction in the risk of ICU admission obtained when vaccinating older adults is nearly equivalent to what obtained by vaccinating rotational workers.

## Discussion

Most studies on vaccine priorities found that an efficient strategy to reduce severe illness due to COVID-19 is to first vaccinate groups that are likely to experience severe illness (Bubar et al., 2021; Babus et al., 2020; Jentsch et al., 2020; Buckner et al., 2020). However, these studies did not consider the impact of disease importations on the infection dynamics. Importations can constitute a significant infection source in regions such as the Atlantic Provinces and the Territories, where most COVID-19 cases are travel-related. We consider both community spread and virus importations by travelers to evaluate the impact of vaccinating rotational workers on reducing the risk of hospitalization and ICU admission. We show that, when disease prevalence at the time of vaccination is low, community spread can be significantly reduced by vaccinating individuals that are likely to introduce the disease in the community, thereby indirectly protecting vulnerable individuals from infection.

We compare the expected number of hospitalizations and ICU admission over four months in the absence of vaccination to this same quantity if rotational workers or vulnerable groups had been vaccinated. We found that, when infection prevalence at the time of vaccination is zero, an X% decrease in the importation rate and an X% decrease in the average probability of hospitalization (or ICU admission) reduce the risk of hospitalization (and ICU admission) in the community by equal amounts. However, if the number of rotational workers is low, reducing disease importations would require significantly less vaccines than reducing the average probabilities of hospitalization and ICU admission. We show that the reduction in the risk of hospitalization and ICU admission when vaccinating 6,000 rotational workers (or 1% of the NL population) can be 3-7 times larger than that obtained if the same number of vaccines were directly given to individuals aged 80 years and older instead. Indeed, vaccinating a small number of older adults would not be sufficient to significantly lower the average hospitalization and ICU admission probabilities amongst the remaining susceptible population when the number of elderly individuals vaccinated is too few. This strategy, however, relies on being able to correctly identify rotational workers, so as to not vaccinate individuals who falsely identify themselves as such.

Vaccinating rotational workers becomes less desirable when infection prevalence in the community at the time of vaccination is high. Our findings are in agreement with the analysis of Russell et al. (2021), showing that imported cases greatly contribute to local epidemic outbreaks in countries with low COVID-19 prevalence, but not in regions with high prevalence. Our analysis shows that the prioritization of rotational workers for vaccination will reduce the importation rate and is likely to prevent community outbreaks that might otherwise have occurred, but we note that it will not be possible to observe these averted community outbreaks. Therefore, while the prioritization of rotational workers should be considered for communities with low infection prevalence to protect their vulnerable groups, the effect of this vaccination strategy can only be assessed with scenario modelling.

Our work provides an analytical criteria that can be easily adopted by public health officials to evaluate whether protection of vulnerable individuals can be achieved by prioritizing members of the mobile labour force for vaccination. Our criteria relates basic elements, such as infection prevalence at the time of vaccination, importation rate, viral transmission after vaccination, and population age structure, to the risk of hospitalization and ICU admission before and after vaccination. Our simple model disentangles the basic relationships between the importation rate and the consequent community infection dynamics, to capture features relevant for planning vaccines prioritization strategies. The analysis presented here can be easily applied to compare vaccinating rotational workers and any vulnerable group known to be at high risk from COVID-19, consisting for example of individuals with health conditions or individuals of other age groups, provided that their vaccination leads to a reduction in the average probabilities of hospitalization and ICU admission of the susceptible population.

For more accurate predictions, future research should consider rotational worker compliance with quarantine, self-isolation and testing requirements (Arino et al., 2020), disease prevalence at work location (affecting the importation rate) (Lopez et al., 2016), the risk of importing new variants of concern (Du et al., 2021), waning immunity (Bubar et al., 2021), non-pharmaceutical interventions, population heterogeneity, and, more generally, stochasticity. Variants of concern are particularly relevant to consider, as immediately after their identification, the prevalence of the particular variant will be low in all other regions. Therefore, our results may apply to all regions of the world with a substantial mobile work force when variants of concern with significant novel effects emerge. Additionally, the dichotomy we consider is to illustrate the value of vaccinating rotational workers, which has been under-explored, however, in practice a multi-pronged approach where vulnerable groups and rotational workers are vaccinated simultaneously is sensible, particularly as more vaccines arrive. Finally, the focus of our work is epidemiological, although economic implications, moral and ethical considerations, and consequences for mental health should also be considered (Giubilini, 2019; Premji, 2020).

## Data Availability

The manuscript contains a mathematical analysis and all data values are provided in the text.

## Acknowledgement

We acknowledge Randy Giffen for suggesting that we conduct this study. AH is supported by grants from the National Science and Engineering Research Council (Discovery Grant), the Atlantic Association for Research in the Mathematical Sciences Centre for Disease Modelling, and the Newfoundland and Labrador Centre for Health Information. AH and MM acknowledge financial support from CANMOD (Canadian Network for Modelling Infectious Diseases - Réseau canadien de modélisation des maladies infectieuses).

## Appendix A: Supplementary Data

### Population

- Age groups considered:

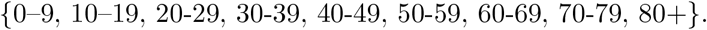
- Population of Newfoundland and Labrador by age group (Population and Demographics):

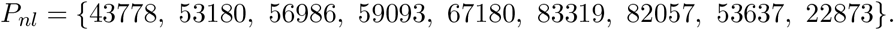
- Proportion of the Newfoundland and Labrador population in each age group (where the total population size *N* = 522, 103 (Population and Demographics)):

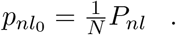
- Proportion of the susceptible Newfoundland and Labrador population in each age group (if individuals aged 85 and above, 10,256 people (Population and Demographics), are vaccinated. We assume that vaccination reduces viral transmission by 0.67% (Knoll and Wonodi, 2021)):

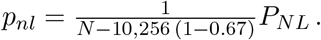
- Proportion of the unvaccinated Newfoundland and Labrador population in each age group (if half of the 80-84 age group, 6,309 people, and individuals aged 85 and above, 10,256 people, are vaccinated. We assume that vaccination reduces viral transmission by 67% (Knoll and Wonodi, 2021)):

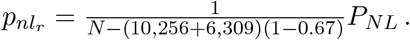

### Probabilities of hospitalization and ICU admission

- Age-stratified probabilities of hospitalization given symptomatic infection (Ferguson et al., 2020):

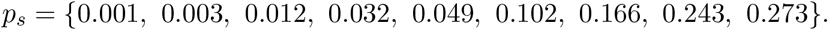
- Age-stratified probabilities of asymptomatic infection (adapted from Kronbichler et al. (2020)):

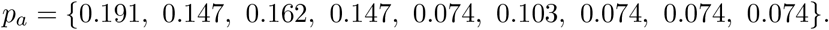
- Age-stratified probabilities of hospitalization (assuming that asymptomatic infections do not result in hospitalization.) (*p*_*h*0_ = *p*_*s*_(1 − *p*_*a*_)):

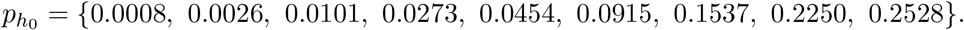
- Age-stratified probabilities of ICU admission given hospitalization (Ferguson et al., 2020):

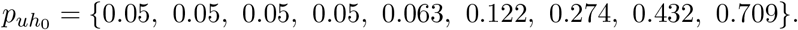
- Age-stratified probabilities of ICU admission (*p*_*u*0_ = *p*_*h*0_ *p*_*uh*0_):

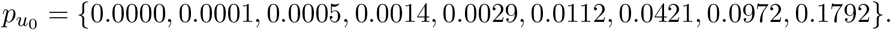

### Probabilities of hospitalization and ICU admission after vaccination of older individuals

To calculate those probabilities we assume that vaccination prevents severe illness (Voysey et al., 2021; Knoll and Wonodi, 2021), and that the probability of hospitalization and ICU admission of vaccinated individuals is 0. In NL, there is 10,256 individuals aged 85 and above, and 12,617 are between 80 and 84 years old (Population and Demographics). Vaccinating half of the 80-84 age group corresponds therefore to vaccinating 6,309 individuals.

- Age-stratified probabilities of hospitalization given infection if individuals aged 85 and above are vaccinated:

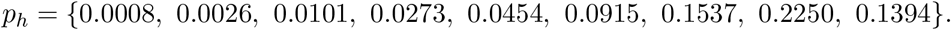
- Average probability of hospitalization if individuals aged 85 and above are vaccinated:

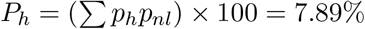
- Age-stratified probabilities of ICU admission if individuals aged 85 and above are vaccinated:

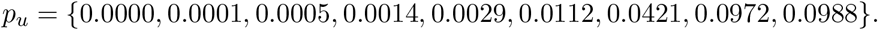
- Average probability of hospitalization if individuals aged 85 and above are vaccinated:

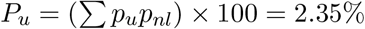
- Age-stratified probabilities of hospitalization if half of the 80-84 age group and individuals aged 85 and above are vaccinated:

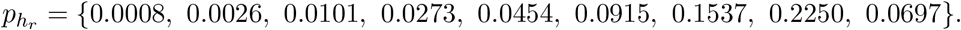
- Average probability of hospitalization when half of the 80-84 age group and individuals aged 85 and above are vaccinated:

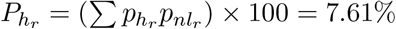
- Age-stratified probabilities of ICU admission if half of the 80-84 age group and individuals aged 85 and above are vaccinated:

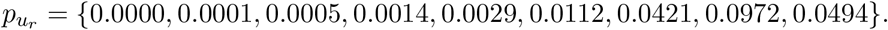
- Average probability of ICU admission if half of the 80-84 age group and individuals aged 85 and above are vaccinated:

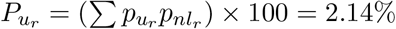
- Most of the rotational workers of Newfoundland and Labrador are between the ages of 25 and 39 (Hewitt et al., 2018). As the number of individuals in those age classes is high, and as the probabilities of hospitalization and ICU admission for those age classes are low, we assume that vaccinating rotational workers does not affect the average probabilities of hospitalization and ICU admission of the susceptible population.

## Appendix B: The impact of *β* on reducing the risk of hospitalization and ICU admission

**Fig. B.1:**
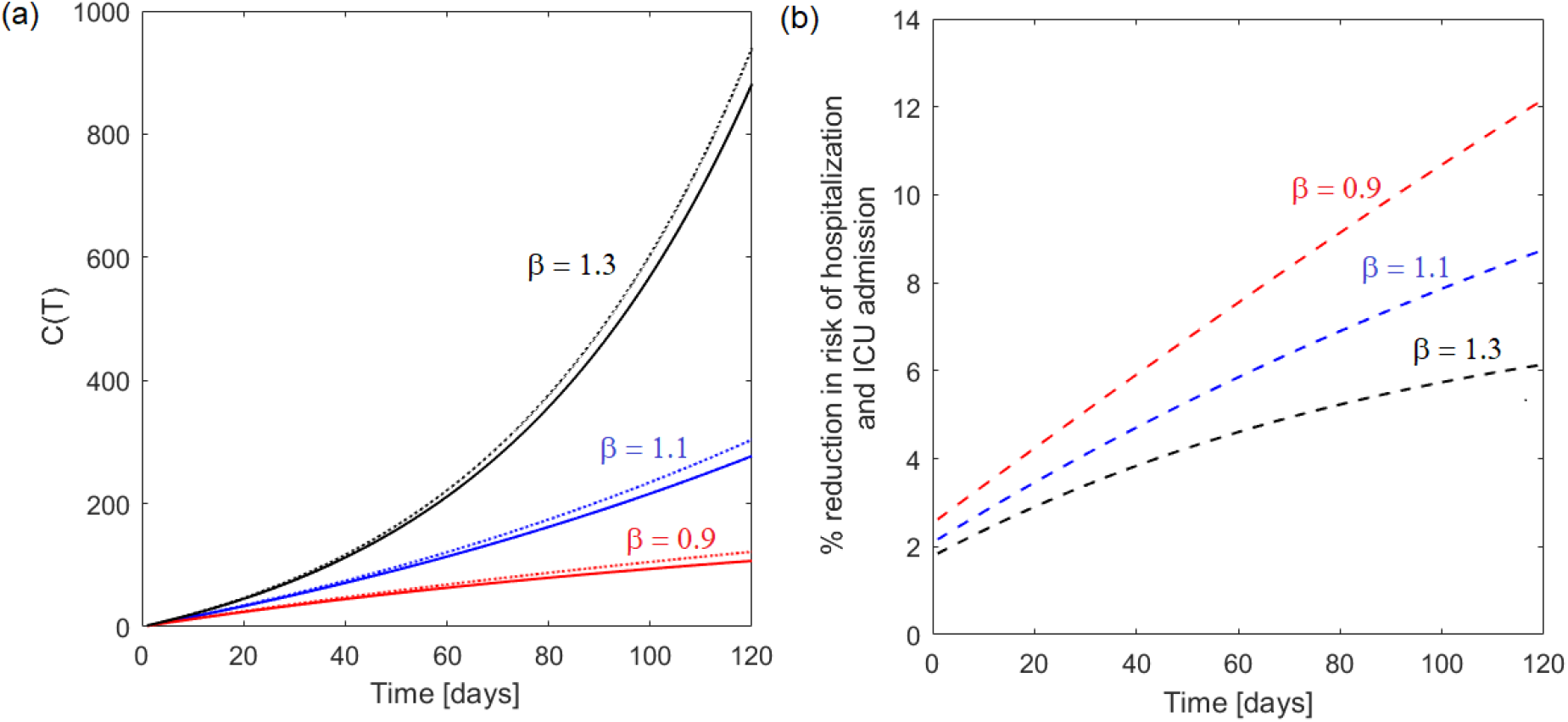
(a) Number of cumulative cases *C*(*T*) as a function of time *T* for different values of the transmission rate *β* (cfr. Eq. (12)). Dotted lines represent *C*(*T*) before a reduction in the importation rate (*m*_0_ = 2/30), while solid lines represent *C*(*T*) after the importation rate have been reduced by 50% (*m*_*r*_ = 1/30). (b) Reduction in the risk of hospitalization and ICU admission due to a reduction in the importation rate, for different values of the transmission rate *β*, where the percent risk reduction is computed as [1 − *C*(*T, m*_0_)*/C*(*T, m*_*r*_)] × 100. In the plots we consider *I*_0_ = 20. For *I*_0_ = 0, the reduction in the risk of hospitalization and ICU admission does not not depend on *β* nor on time *T* (cfr. Eq. (13)) and it is constant at 50% (i.e., [1 − *m*_*r*_ */m*_0_] × 100). For *I*_0_ > 0 the impact of decreasing importations to reduce hospitalization and ICU admission is larger when the transmission rate *β* is low and when longer time intervals are considered.

**Fig. B.2:**
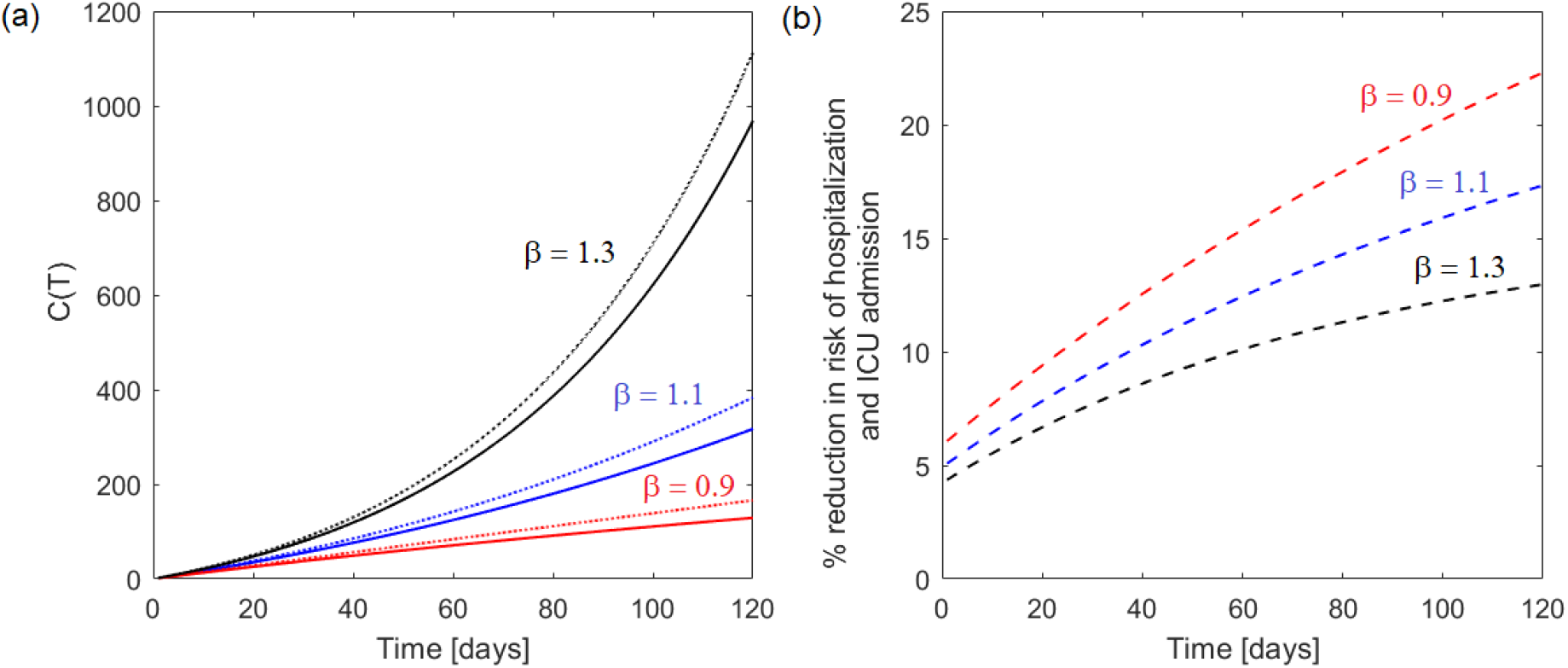
Same as Fig. B.1 but the importation rate is larger, with *m*_0_ = 5/30 and *m*_*r*_ = 2.5/30.

